# Hamstring strain grade on MRI and return to play in elite Australian cricket players

**DOI:** 10.1101/2024.11.07.24316885

**Authors:** Thomas Cooney, Ashton Reeve, Anna E. Saw, Alex Kountouris, John W. Orchard, James Linklater

## Abstract

**Objective:** i) determine whether the grade of hamstring strain confirmed by magnetic resonance imaging (MRI) is related to time to return to play, and ii) describe the incidence, prevalence and grade of hamstring strains confirmed by MRI in elite Australian cricket players.

**Design:** Retrospective case series.

**Methods:** Hamstring strains from professional domestic and international cricket teams over 13.5 seasons which had received MRI scans were graded using British Athletics Muscle Injury Classification (BAMIC) system. The main outcome measure was time to return to play.

**Results:** 141 hamstring strain injuries with available MRI imaging scans were recorded during the study period (average 3.2 per 100 players per season: male 4.5, female 1.2). The most commonly injured muscle was biceps femoris (64%, 95% CI 56-71%) and the most frequent category of injury was grade 2C (27%, 20-35%). Across all injury grades, players were unavailable for full participation for a median of 23 (IQR 15-38) days and missed 3 (1-6) matches. The number of days unavailable were higher for injuries which were graded 2 or 3, compared to grade 1 (p=0.018, p=0.002 respectively), and injuries which included the tendon compared to those which did not (p=0.002).

**Conclusions:** This study provides evidence that higher grade injuries and those involving the intramuscular tendon are associated with a more prolonged return to play. This finding should be viewed in context of the study limitation that clinicians treating players were not blinded to the MRI findings.

## 1. Introduction

Hamstring strain injuries are consistently the most common time-loss injury experienced by elite cricket players.^[1]^ The incidence of hamstring strain injuries which cost match time has been reported as 7.4 and 3.1 new injuries per 100 players per season in elite Australian male and female cricket players respectively.^[1]^ In elite male players from England and Wales, the incidence has been reported as 5.9, ^[2]^ and 3.1 in male players from New Zealand.^[3]^ The higher incidence of hamstring strains in more recent years may reflect a shift to shorter match formats (one-day, T20) from multi-day match formats. ^[2]^

In professional sport, MRI is commonly used both to diagnose and prognosticate acute hamstring injuries. Many different classification systems have been proposed and used to categorise hamstring injuries based on their radiological findings ^[4]^ but debate continues around their utility in predicting times to return to play.^[5]^ The British Athletics Muscle Injury Classification (BAMIC) system classifies muscle strains based on site of injury (myofascial; muscle-tendon junction; intra-tendon) and extent of injury (% of cross-sectional area and cranio-caudal length).^[6]^ The BAMIC system is a reliable and widely used system and its grading has been associated with return to play from hamstring strain injury in professional soccer,^[7]^ Australian Rules Football ^[8]^ and track and field athletics.^[9]^ Reviews of the limited and heterogeneous literature to-date have concluded that there is moderate, yet not universally consistent, evidence for injuries involving the intramuscular tendon to have a prolonged return to play time and an increased risk of re-injury.^[10, 11]^ Additional research on other athletic cohorts such as cricketers will contribute further evidence.

The primary aim of this study was to determine whether the grade of hamstring strain confirmed by MRI in elite Australian male and female cricket players is related to time to return to play. The secondary aim of this study was to describe the incidence, prevalence and grade of hamstring strains confirmed by MRI in elite Australian cricket players. This study hypothesises that higher grade hamstring strains and injuries involving the intramuscular tendon are associated with an increased time to return to play.

## 2. Methods

Ethics approval was granted from La Trobe University Human Ethics Committee (HEC20058) which waived the need for individual informed consent.

Data was retrieved from Cricket Australia’s online Athlete Management System (AMS) (Fair Play AMS Pty Ltd, Australia) for the 13.5 seasons from 1 July 2009 – 31st December 2022 (seasons delineated as 1 July to 30 June). Demographic data were sex, age, dominant skill, and dominant (throwing) hand. Injury data is recorded at the time of injury in a standardised manner by medical staff (doctor and/or physiotherapist) working with state/territory and national cricket teams.

### Injury data

Hamstring strain injuries were included if: they occurred during a cricket match, training, conditioning or warm-up; they were diagnosed by the treating doctor/physiotherapist at the time as an acute hamstring muscle or tendon injury, and; the diagnosis was confirmed by MRI within 21 days of injury. Injuries were excluded if: they occurred outside of an organised cricket activity (e.g., other sport, movement at home); they were diagnosed by the treating doctor/physiotherapist as tendinopathy, neural hamstring pain or an overuse injury, or; there was no MRI available for review.

If players had a history of hamstring strain, the injury was considered recurrent if it occurred on the same side as a previous injury. Recurrent injuries were further classified by the time between returning to full participation from the index injury and the occurrence of the recurrent injury: early (within 2 months), late (2-12 months), delayed (more than 12 months).^[12]^

Return to full training/match availability was determined from one or more of: the treating doctor/physiotherapist deemed the player available to play a match (regardless of if a match were on that day); clinical notes stated the player may return to unrestricted training/match participation, or; the player participated in a match. If an accurate date of return to full training/match availability could not be determined retrospectively, this was left blank.

### MRI data

MRI data was viewed on Inteleviewer (Intelerad, Montreal, Canada). Following a training period, two authors (TC (Sport and Exercise Medicine Registrar) and AR (Musculoskeletal Radiologist)) independently reviewed all images and graded hamstring strains according to the British Athletics Muscle Injury Classification (BAMIC). ^[6]^ Data between reviewers was compared and any discrepancies in grading were adjudicated by a senior radiologist (JL). The researchers reviewing the MRI were blinded to clinical information including individual characteristics, diagnosis, and injury history to ensure MRI were graded objectively. Researchers were also blinded to the radiology report.

The BAMIC includes a subjective rating of both the grade (size) of the injury and location of the injury.^[6]^ Grade 0 is defined by no abnormal findings on MRI; grade 1 if oedema extends < 10% of the muscle cross sectional area (CSA) or <5cm of the longitudinal length with <1cm fibre disruption; grade 2 if 10-50% CSA or 5-15cm length with <5cm disruption; grade 3 if >50% CSA or >15cm length or >5cm disruption, and; grade 4 if a complete tear. Grades 1-3 are subclassified by locations A-C, where A is myofascial, B is muscle or myotendinous junction, and C is tendinous.

### Analyses

Descriptive statistics were used to characterise the players included in the sample and the presentation and circumstances of injuries. Injury incidence was calculated as the number of injuries per 100 players per season. Average injury incidence was calculated as the average of each incidence per season. Proportions were calculated as a percentage of the number of data points known for each variable, with a Wilson 95% confidence interval (CI). Days unavailable and matches missed were not normally distributed, hence descriptive statistics are reported as median and interquartile range (IQR). Median tests and independent-samples Kruskal-Wallis tests were used to compare between BAMIC grades in injury locations. A generalised estimating equation with an exchangeable correlation structure was used to examine the linear relationship between age and days unavailable, accounting for individuals with multiple injuries and controlling for sex as a cofactor. Injuries with an unsuccessful return to full training/match availability were excluded from the group analyses for return to play statistics and investigated separately. Additional analyses separated pace bowlers from the other dominant skills to determine if return to play statistics differed given the additional demands of pace bowling on the hamstrings. Analyses were conducted in Excel (Microsoft, 2016 MSO) and SPSS (version 28, IBM, Armonk, NY, USA).

## 3. Results

The study population included 478 male and 365 female cricket players who were contracted during the study period (average of 174 and 121 players per season respectively).

141 instances of hamstring strain met the inclusion criteria over the 13.5 seasons (male n=119, mean age 27.9 ± 4.4 years; female n=22, mean age 26.5 ± 5.4 years). The distribution of injury grades and return to play times were similar between males and females, and the number of female injuries too few for sub-analysis, male and female data are analysed collectively henceforth. The average incidence of hamstring strains confirmed by MRI over the 13.5 seasons was 10.1 injury instances per season (3.2 per 100 players per season: male 4.5, female 1.2, p<0.001 comparing male and female incidence across each season). The incidence varied across seasons as depicted in Figure 1. Batters (45%, 95% CI 37-54%), pace bowlers (33%, 25-41%), and all-rounder pace bowlers (11%, 7-17%) sustained the majority of injuries.

**Figure 1.**
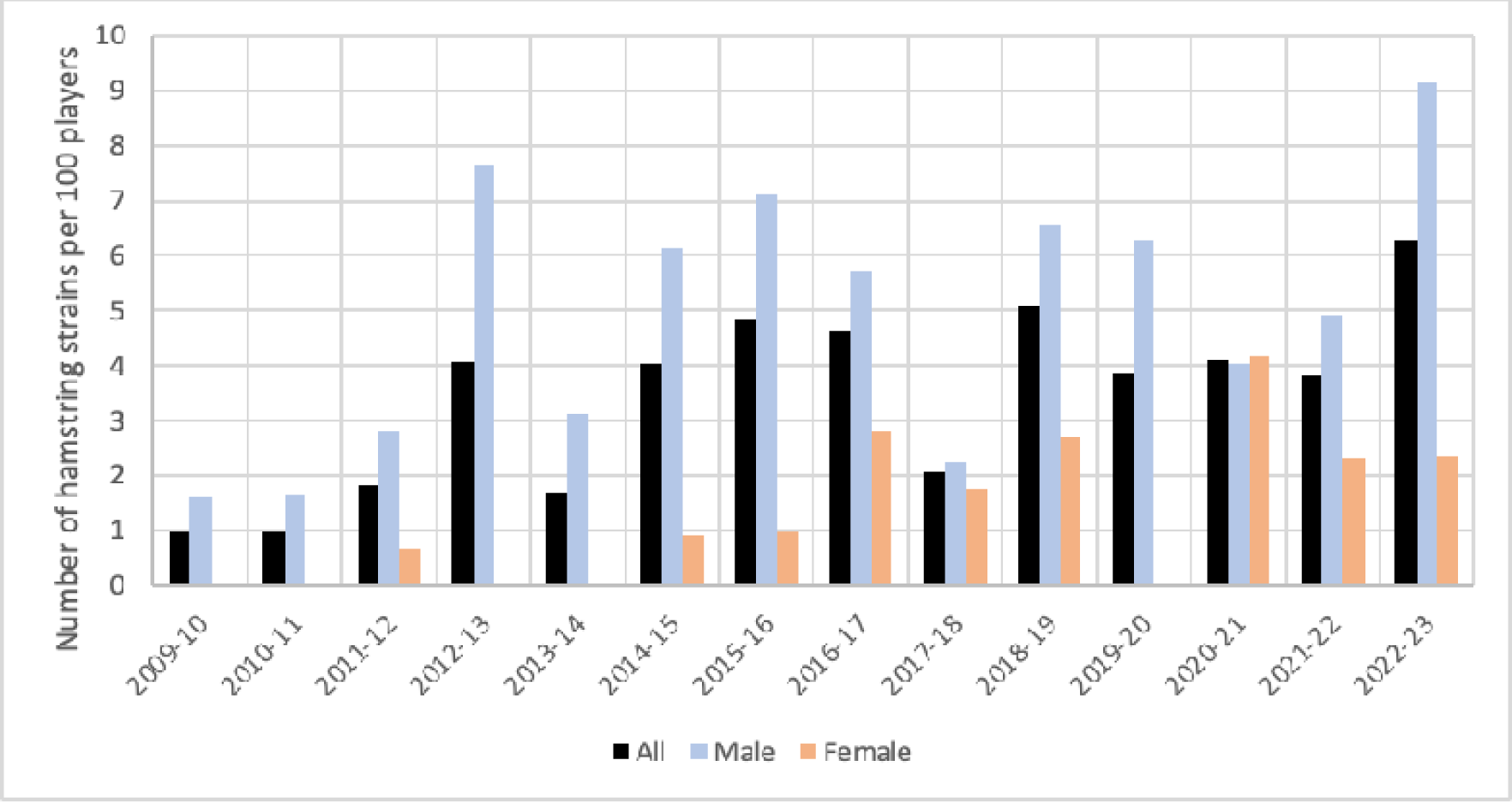
Incidence of hamstring strains confirmed by MRI in elite Australian cricket players over 13.5 seasons.

Injuries were sustained during a match (68%, 60-75%), cricket conditioning (17%, 12-24%), or cricket training (14%, 9-21%). Match formats where injuries occurred were 20-over (42%, 32-52%), one-day (29%, 20-39%), and multi-day (30%, 21-40%).

Injuries sustained during a match occurred whilst batting (including running between wickets) (41%, 32-51%), bowling (31%, 22-40%), and fielding (28%, 20-38%). Running was the most common mechanism (62%, 54-70%), followed by bowling (23%, 17-31%), fielding the ball by bending, lunging, diving, and/or changing direction (11%, 7-17%), and batting (4%, 2-8%).

Biceps femoris was the most commonly injured muscle (64%, 56-71%), followed by semitendinosus (11%, 7-17%) and semimembranosus (8%, 4-13%). In 18% (12-25%) of cases, no muscle was specified due to a lack of findings on imaging. The injured side relative to the dominant throwing hand was contralateral (53%, 45-61%) or ipsilateral (47%, 39-55%).

The BAMIC grading of injuries are detailed in Table 1. Across all injuries, players were unavailable for full participation for a median of 23 (IQR 15-38) days and missed 3 (1-6) matches. The number of days unavailable were higher for injuries which were graded 2 or 3, compared to grade 1 (p=0.018, p=0.002 respectively), and for injuries which included the tendon (C) compared to those which did not (B; p=0.002) (Table 2, Figure 2). Pace bowlers had a higher number of days unavailable per injury for injuries which involved the muscle or myotendinous junction (B), or tendon (C) compared to players with other dominant skills (p=0.060). Age was not related to the number of days unavailable (p=0.222).

**Table 1.** Proportion of hamstring strain injuries sustained by elite Australian cricket players categorised by the British Athletics Muscle Injury Classification (BAMIC). Proportion presented as percentage with 95% confidence interval.

| BAMIC Grade | Proportion of all injuries (n=141) |
| --- | --- |
| 0 | 18 (12-25) |
| 1A | 6 (3-12) |
| 1B | 3 (1-7) |
| 2A | 19 (14-26) |
| 2B | 16 (11-23) |
| 2C | 27 (20-35) |
| 3A | 0 (0-3) |
| 3B | 0 (0-3) |
| 3C | 8 (4-13) |
| 4 | 0 (0-3) |
| 4C | 3 (1-7) |

**Table 2.**
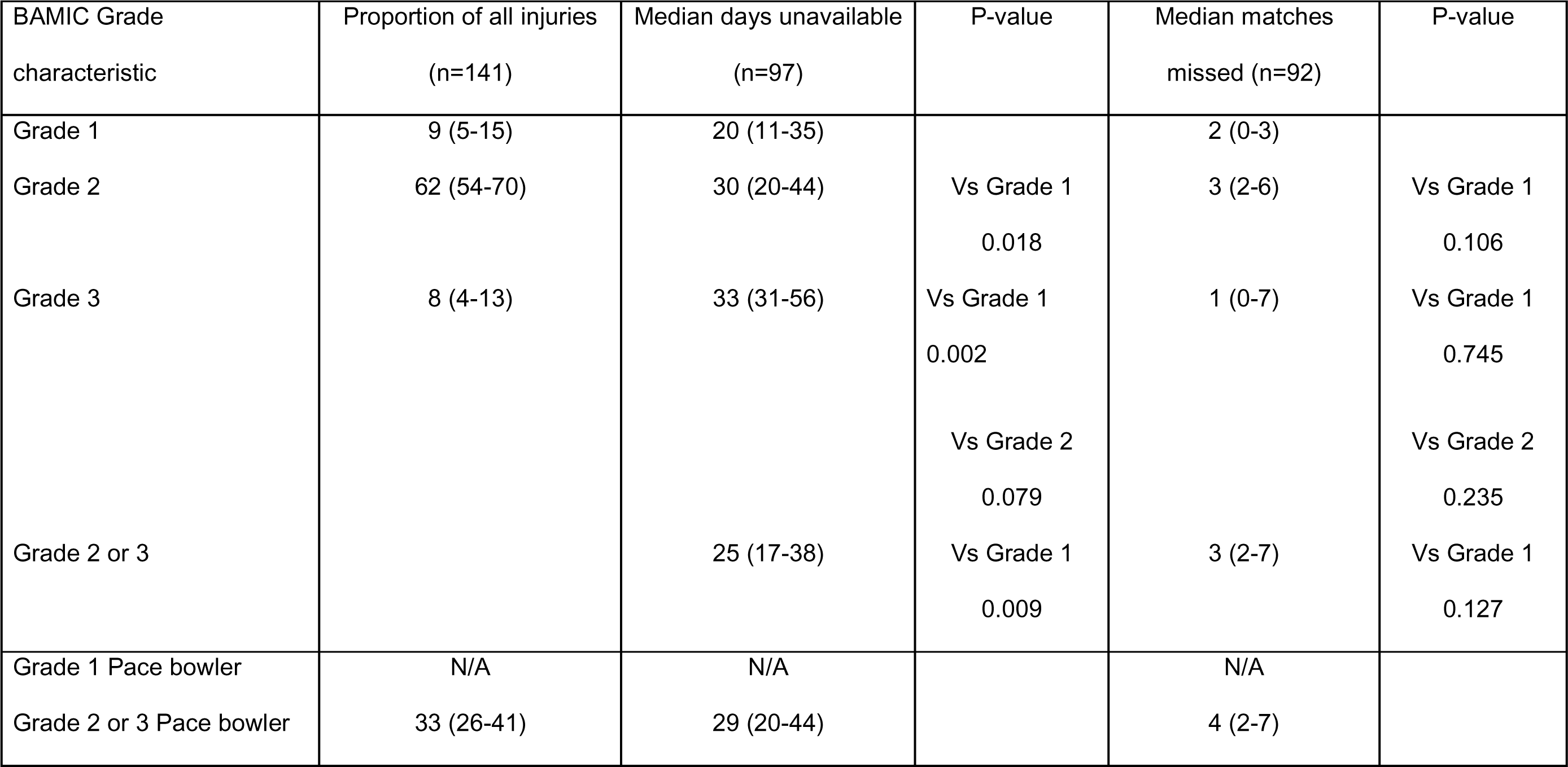
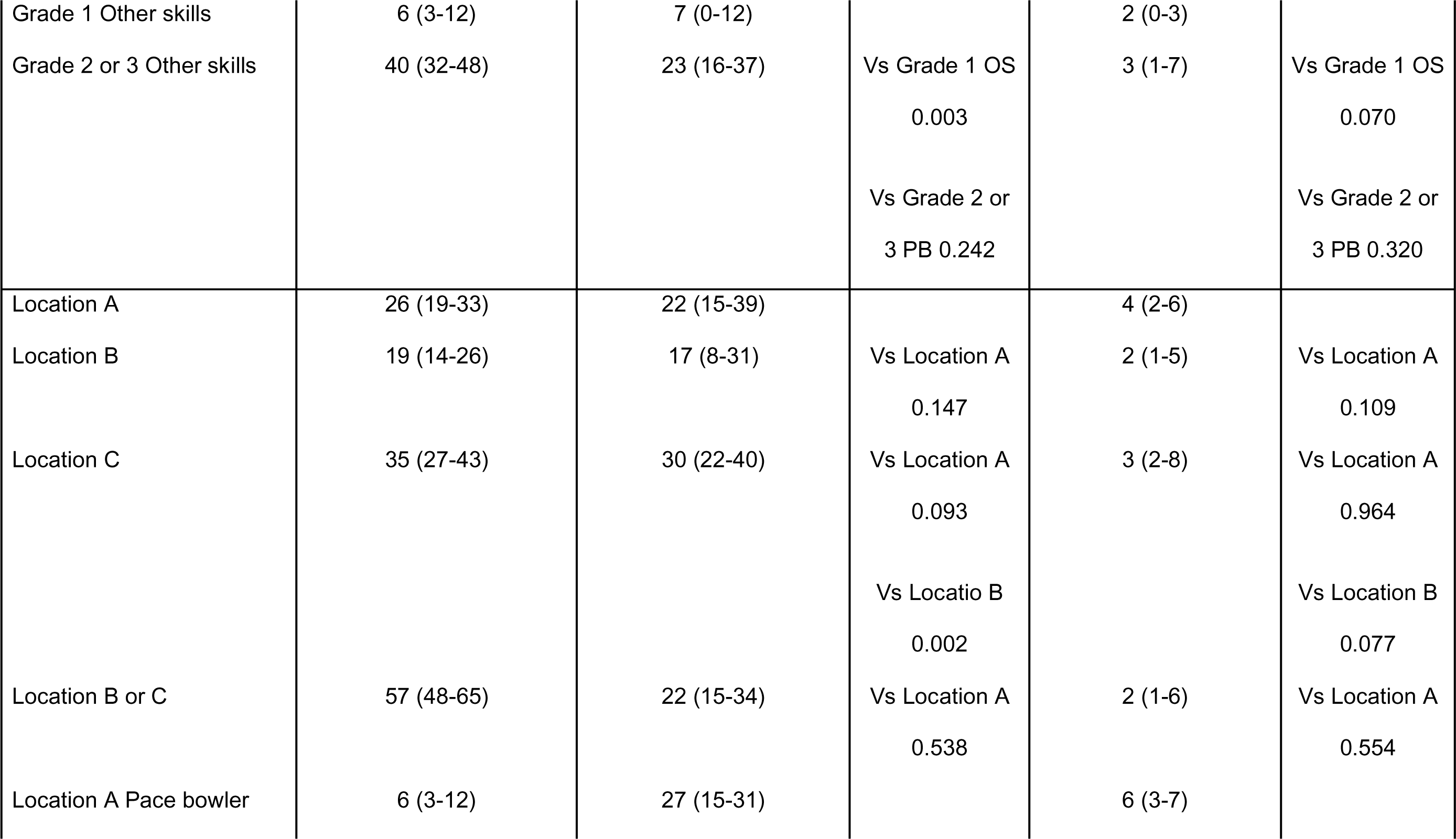
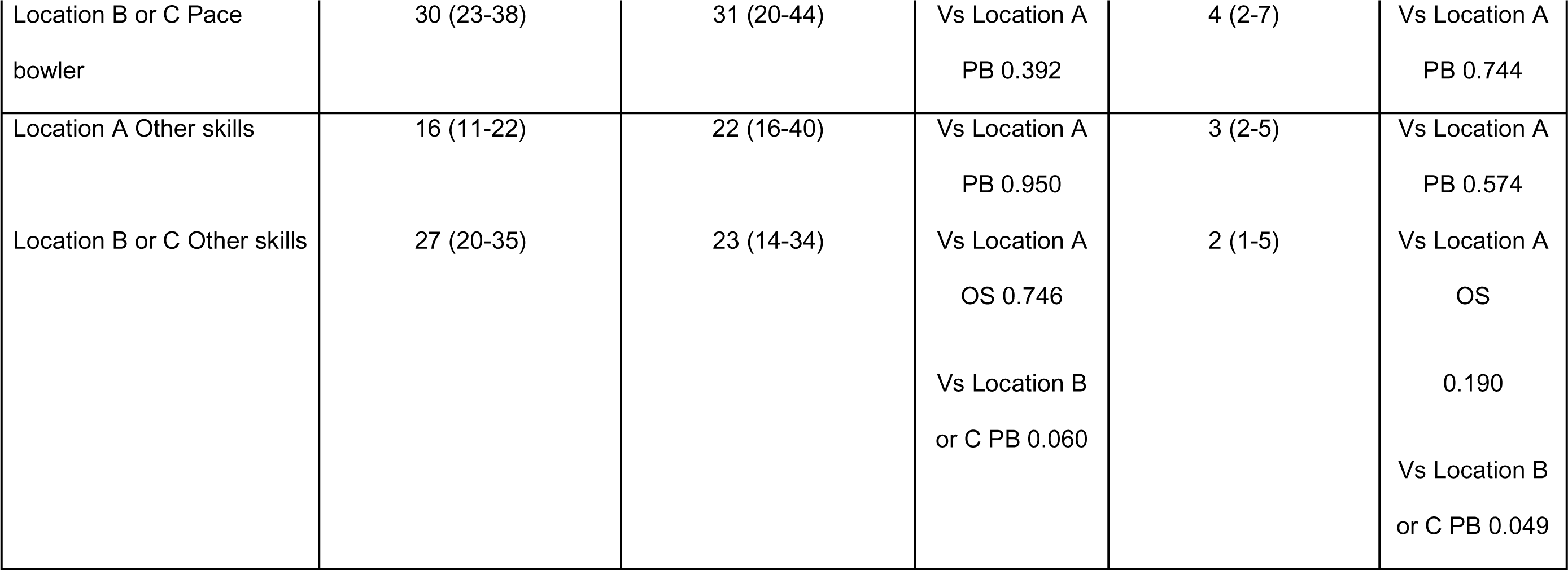
Days unavailable and matches missed due to hamstring strain injuries categorised by size and tendon involvement as per the British Athletics Muscle Injury Classification (BAMIC). Proportion presented as percentage with 95% confidence interval and median presented with interquartile range. PB = Pace bowler, OS = Other skills. N/A if n<5.

Twenty-six recurrent injuries were observed: 4 early, 5 late, and 17 delayed recurrences. In 21 of these recurrences (81%, 62-91%), the specific muscle involved was the same as that of the index injury. Of these 21 recurrences, the BAMIC grading of injury was more severe in 6 injuries, the same in 13 injuries, and less severe in 2 injuries.

## 4. Discussion

This study analysed 13.5 seasons of data from elite Australian cricket players, with a steady increase in the incidence of hamstring strain injury from the beginning of the study period to the end of 2022. There was an average incidence of 3.2 hamstring strains per 100 players per season which were confirmed by MRI, with a higher incidence in males than females. This may in part be due to the male season traditionally being longer, with more training sessions per week and more matches played per calendar year.^[13]^ Similar trends have been seen in soccer,^[14]^ athletics ^[15]^ and field hockey.^[15]^ Sex-based differences which may contribute to male athletes being 2-4 times more likely to sustain a hamstring injury compared with females include differences in hamstring musculotendinous stiffness, muscle endurance and hamstring to quadricep strength ratio.^[16]^

Hamstring injuries were more likely to occur in a match compared with training or conditioning sessions. This is likely due to the increased intensity and pressure of matches relative to training and the increased likelihood of performing under fatigue. T20 cricket was associated with the highest risk of hamstring strain injury, in keeping with the known increase in intensive movements and high-speed running required in this format.^[17, 18]^

Batters running between the wickets was the most common action during which a hamstring injury occurred. Previous studies have found in shorter formats running between the wickets is the most common mechanism,^[1, 2, 19, 20]^ whereas in multi-day formats, fast bowling is a more common injury mechanism.^[2]^ There are a number of possibilities why batting injuries are common, including the intensity of running and the need to decelerate, change direction and rapidly accelerate again in a fatigued state when scoring (running) multiple runs. Players accelerate and decelerate with the hamstrings in a lengthened position when stretching out to ground their bat which could put them at further risk of strain injury. Batters also run while wearing pads and carrying their bat which may play a role in changing their normal running mechanics.

Pace bowlers had a higher number of days unavailable for injuries involving the muscle or myotendinous junction (B) and the intramuscular tendon (C) compared to players with other dominant skills. This is likely due to the physical demands of pace bowling, which includes a high speed run up, a pre-delivery stride and release of the ball followed by rapid deceleration and a follow through.^[21]^ This information may assist clinicians in their decision making around returning to play with pace bowlers who have been diagnosed with a strain which involves the myotendinous junction or intramuscular tendon.

Biceps femoris was the most commonly injured muscle, accounting for 63% of all hamstring injuries. This has been observed in other sports involving sprinting, including soccer,^[22]^ Australian Rules Football ^[23]^ and athletics.^[24]^ The exact reason for this observation is as yet unknown, however a number of reasons have been hypothesised, including the dual innervation of the biceps femoris, the increased number of type II muscle fibres with the biceps femoris and the biceps being required to exert more force relative to the two other hamstring muscles during a lengthening muscle contraction.^[25, 26]^

More extensive injuries (Grade 3 and 4) and those involving the intramuscular tendon or free tendon (C) were found to be related to the number of days unavailable to train or play. Similar results have been found in studies of athletes across multiple sports,^[11]^ including AFL,^[8]^ Track and field ^[27]^ and soccer.^[7]^ According to James et al, tendon healing occurs at a much slower rate when compared with muscle healing. This is due to the prolonged time required for the synthesis of Type 1 collagen and for tendon remodelling to occur, a process which begins 6 weeks after the initial injury.^[28]^ Adequate remodelling is required for the tendon to withstand the tensile loads required during sport specific activities like sprinting and kicking.

Wangensteen et al found similar results in their 2016 study, reporting that 79% of injuries occurred in the same location within the muscle as the index injury.^[29]^ Contrastingly however, they found that all re-injuries occurring in the same location as the index injury were more severe based on radiological grading. In Australian Rules Football, recurrences of muscle strains have decreased from the era in the 1990s (where MRI scans were rarely used) to the modern era (where the vast majority of strains receive an MRI).^[30]^ It may be that MRI scans promote conservatism of management which leads to decreased recurrences.

This study has a number of strengths. Data was collected over a long period of time (13.5 seasons) and included a large cohort of patients. All MRI were read consistently by two researchers experienced in reading musculoskeletal MRI and grading injuries according to BAMIC. Researchers grading the MRI were blind to clinical information and were not involved with the diagnosis and management of the cricket players during the study period. The retrospective analysis of clinical data means that the medical staff were not influenced by the study.

An important limitation of this retrospective study is that clinicians used the MRI to inform their management of injuries at the time, and therefore may have been more conservative for players with higher graded injuries. The findings of this study relating injury grade to return to play times must be viewed in the context of this limitation. Additional limitations to note are that the clinical management of players was performed by a number of different practitioners over a number of years across differing states and territories. Therefore, there may be inconsistencies in the ordering of imaging and the rehabilitation of injuries. This in turn has the potential to impact the inclusion criteria and the return to play data. A further limitation is that group-level analyses do not capture the nuance of individual situations, particularly in elite sport. For example, the decision to manage a player a certain way may be influenced by the time of season, the importance of upcoming matches, and other individual factors and preferences.

## 5. Conclusion

Hamstring injuries are common in elite level cricket. The use of MRI to diagnose and grade hamstring injuries is now widespread in professional sport, with the BAMIC system one of the most commonly used methods of grading MRI. This study provides further evidence that higher grade injuries and those involving the intramuscular tendon are associated with a more prolonged return to play. Pace bowlers, in particular, had a longer time to return to play compared with non-pace bowlers when there was involvement of the musculotendinous junction or the intramuscular tendon. This study provides useful information for clinicians managing injuries in elite cricket players, particularly those rehabilitating hamstring strains in pace bowlers.

## 6. Practical implications

- Approximately three percent of cricket players sustain a hamstring strain each season, with higher-risk activities being running between the wickets while batting, pace bowling, and fielding during matches.
- More extensive injuries (Grade 3 and 4) and those involving the intramuscular tendon or free tendon (C) were found to have prolonged return to play times, a finding consistent with other sports. Return to play time was particularly prolonged in pace bowlers.
- Medical staff should consider the grade of hamstring strain on MRI and the specific physical demands required of a player when managing the return to play of an injured player.

## Data Availability

All data produced in the present study are available upon reasonable request to the authors

## Confirmation of Ethical Compliance

Ethics approval was granted from La Trobe University Human Ethics Committee (HEC20058)

## CRediT authorship contribution statement

TC: Conceptualization, methodology, formal analysis, investigation (MRI reading), data curation, writing – original draft, writing – review and editing, project administration.

AR: Formal analysis, investigation, data curation, (MRI reading)

AS: Conceptualization, methodology, writing – original draft, project administration, writing – review and editing

AK: Conceptualization, methodology, writing – review and editing

JO Conceptualization, methodology, writing – review and editing, corresponding author

JL: Formal analysis, investigation (MRI reading)

## Acknowledgements

The authors acknowledge the doctors and physiotherapists working for Australian cricket teams who recorded the clinical data and radiology practices from around Australia who conducted the imaging.

## References

1. Orchard JW, Inge P, Sims K, et al. Comparison of injury profiles between elite Australian male and female cricket players. J Sci Med Sport. 2023; 26(1):19–24.

2. Goggins L, Langley B, Griffin S, et al. Hamstring injuries in England and Wales elite men’s domestic cricket from 2010 to 2019. J Sci Med Sport. 2022; 25(6):474–479.

3. Frost WL, Chalmers DJ. Injury in elite New Zealand cricketers 2002-2008: descriptive epidemiology. Br J Sports Med. 2014; 48(12):1002–1007.

4. Wangensteen A, Guermazi A, Tol JL, et al. New MRI muscle classification systems and associations with return to sport after acute hamstring injuries: a prospective study. Eur Radiol. 2018; 28(8):3532–3541.

5. Hamilton B, Alonso JM, Best TM. Time for a paradigm shift in the classification of muscle injuries. J Sport Health Sci. 2017; 6(3):255–261.

6. Pollock N, James SL, Lee JC, Chakraverty R. British athletics muscle injury classification: a new grading system. Br J Sports Med. 2014; 48(18):1347–1351.

7. Shamji R, James SLJ, Botchu R, Khurniawan KA, Bhogal G, Rushton A. Association of the British Athletic Muscle Injury Classification and anatomic location with return to full training and reinjury following hamstring injury in elite football. BMJ Open Sport Exerc Med. 2021; 7(2):e001010.

8. Eggleston L, McMeniman M, Engstrom C. High-grade intramuscular tendon disruption in acute hamstring injury and return to play in Australian Football players. Scand J Med Sci Sports. 2020; 30(6):1073–1082.

9. Macdonald B, McAleer S, Kelly S, Chakraverty R, Johnston M, Pollock N. Hamstring rehabilitation in elite track and field athletes: applying the British Athletics Muscle Injury Classification in clinical practice. Br J Sports Med. 2019; 53(23):1464–1473.

10. Beattie CE, Barnett RJ, Williams J, Sim J, Pullinger SA. Are return-to-play times longer in lower-limb muscle injuries involving the intramuscular tendon? A systematic review. J Sci Med Sport. 2023; 26(11):599–609.

11. Kerin F, O’Flanagan S, Coyle J, et al. Intramuscular Tendon Injuries of the Hamstring Muscles: A More Severe Variant? A Narrative Review. Sports Med Open. 2023; 9(1):75.

12. Fuller CW, Ekstrand J, Junge A, et al. Consensus statement on injury definitions and data collection procedures in studies of football (soccer) injuries. Scand J Med Sci Sports. 2006; 16(2):83–92.

13. Panagodage Perera NK, Kountouris A, Kemp JL, Joseph C, Finch CF. The incidence, prevalence, nature, severity and mechanisms of injury in elite female cricketers: A prospective cohort study. J Sci Med Sport. 2019; 22(9):1014–1020.

14. Cross KM, Gurka KK, Saliba S, Conaway M, Hertel J. Comparison of hamstring strain injury rates between male and female intercollegiate soccer athletes. Am J Sports Med. 2013; 41(4):742–748.

15. Cross KM, Gurka KK, Conaway M, Ingersoll CD. Hamstring strain incidence between genders and sports in NCAA Athletics. Athletic Training & Sports Health Care. 2010; 2(3):124–130.

16. O’Sullivan L, Tanaka M. Sex-based differences in hamstring injury risk factors. Journal of Women’s Sports Medicine. 2021; 1(1):20–29.

17. Orchard J, James T, Kountouris A, Portus M. Changes to injury profile (and recommended cricket injury definitions) based on the increased frequency of Twenty20 cricket matches. Open Access J Sports Med. 2010; 1:63–76.

18. Petersen CJ, Pyne D, Dawson B, Portus M, Kellett A. Movement patterns in cricket vary by both position and game format. J Sports Sci. 2010; 28(1):45–52.

19. Orchard JW, Kountouris A, Sims K. Risk factors for hamstring injuries in Australian male professional cricket players. J Sport Health Sci. 2017; 6(3):271–274.

20. Goggins L, Williams S, Griffin S, Langley B, Newman D, Peirce N. English and Welsh men’s domestic cricket injury risk by activity and cricket type: A retrospective cohort study from 2010 to 2019. J Sci Med Sport. 2024; 27(1):25–29.

21. Bartlett RM, Stockill NP, Elliott BC, Burnett AF. The biomechanics of fast bowling in men’s cricket: a review. J Sports Sci. 1996; 14(5):403–424.

22. Crema MD, Guermazi A, Tol JL, Niu J, Hamilton B, Roemer FW. Acute hamstring injury in football players: Association between anatomical location and extent of injury-A large single-center MRI report. J Sci Med Sport. 2016; 19(4):317–322.

23. Slavotinek JP, Verrall GM, Fon GT. Hamstring injury in athletes: using MR imaging measurements to compare extent of muscle injury with amount of time lost from competition. AJR Am J Roentgenol. 2002; 179(6):1621–1628.

24. Askling CM, Tengvar M, Saartok T, Thorstensson A. Acute first-time hamstring strains during high-speed running: a longitudinal study including clinical and magnetic resonance imaging findings. Am J Sports Med. 2007; 35(2):197–206.

25. Dolman B, Verrall G, Reid I. Physical principles demonstrate that the biceps femoris muscle relative to the other hamstring muscles exerts the most force: implications for hamstring muscle strain injuries. Muscles Ligaments Tendons J. 2014; 4(3):371–377.

26. Opar DA, Williams MD, Shield AJ. Hamstring strain injuries: factors that lead to injury and re-injury. Sports Med. 2012; 42(3):209–226.

27. Pollock N, Kelly S, Lee J, et al. A 4-year study of hamstring injury outcomes in elite track and field using the British Athletics rehabilitation approach. Br J Sports Med. 2022; 56(5):257–263.

28. James R, Kesturu G, Balian G, Chhabra AB. Tendon: biology, biomechanics, repair, growth factors, and evolving treatment options. J Hand Surg Am. 2008; 33(1):102–112.

29. Wangensteen A, Tol JL, Witvrouw E, et al. Hamstring Reinjuries Occur at the Same Location and Early After Return to Sport: A Descriptive Study of MRI-Confirmed Reinjuries. Am J Sports Med. 2016; 44(8):2112–2121.

30. Orchard JW, Chaker Jomaa M, Orchard JJ, et al. Fifteen-week window for recurrent muscle strains in football: a prospective cohort of 3600 muscle strains over 23 years in professional Australian rules football. Br J Sports Med. 2020; 54(18):1103–1107.

